# An optimal lockdown relaxation strategy for minimizing the economic effects of covid–19 outbreak

**DOI:** 10.1101/2020.06.08.20125583

**Authors:** A.C. Mahasinghe, K.K.W.H. Erandi, S.S.N. Perera

## Abstract

In order to recover the damage to the economy by the ongoing covid–19 pandemic, many countries consider the transition from strict lockdowns to partial lockdowns through relaxation of preventive measures. In this work, we propose an optimal lockdown relaxation strategy, which is aimed at minimizing the damage to the economy, while confining the covid–19 incidence to a level endurable by the available healthcare facilities in the country. In order to capture the transmission dynamics, we adopt the compartment models and develop the relevant optimization model, which turns out to be non–linear. We generate approximate solutions to the problem, whereas our experimentation is based on the data on the covid–19 outbreak in Sri Lanka.

## 1 Introduction

The ongoing pandemic of the covid–19, which has recently been declared by the World Health Organization as the era–defining global health crisis [16], is an unprecedented threat to the world. It has caused numerous deaths and health complications, upended the day–to–day life and destabilized economies.

On 23^rd^ January, 2020 the State Council of Wuhan issued the first order to lockdown the first pandemic epicenter to control the spread of the virus to the other parts of the country [26]. However, the virus quickly spread throughout the globe within the first two months of 2020. In the absence of established measures to control the transmission of covid–19, at least 186 countries have implemented various strategies including social distancing and restrictions on human movement to slow down the spread of the disease [10] and to prevent healthcare systems from becoming overwhelmed. For instance, as the first European country to be massively affected, Italy issued a lockdown order in a cluster of cities in Lombardy and Veneto regions on 22^nd^ February, 2020 and it was further expanded to 15 provinces on 8^th^ March; subsequently imposed a nationwide lockdown on 12^th^ March [21]. The government of Philippines issued a lockdown order in the form of an enhanced community quarantine on 12^th^ March, 2020 [20]. The government of France too declared a mandatory home confinement for 15 days starting from 17^th^ March, 2020 which was extended up to 30 days and lifted on 11^th^ May, 2020 [13]. The government of India declared a countrywide lockdown for 21 days on 24^th^ March, 2020; but further extended up to 3^rd^ May, 2020 [14] as the number of covid–19 cases increased.

Though lockdown and other control strategies had been able to control the spread of the disease and saved many lives, most of the countries experienced considerable socio–economic damages during the lockdown period. For instance, the collapse of markets, employee layoffs, disruptions of the food supply and impacts on education system can be highlighted [2,15,19]. To overcome these situations many countries gradually lifted the strict lockdowns. However, the consequence was another epidemic wave. For an example, after the lockdown relaxation on 3^rd^ May 2020, India is witnessing a steep rise in the number of reported covid–19 cases as shown in Figure 1.

**Fig. 1:**
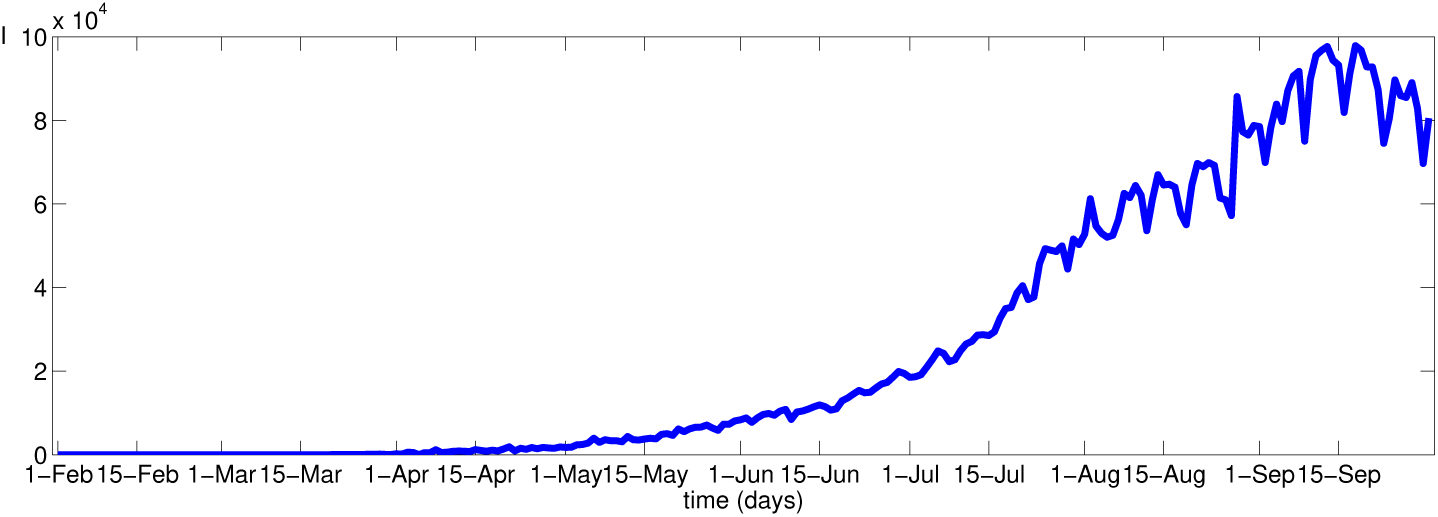
Daily reported COVID–19 cases from 31^st^ January 2020 to 29^th^ September 2020, India (Source: Worldometer)

Figures 2 and 3 clearly illustrate that Italy and France are experiencing the second phase of pandemic after the lockdown relaxation. From Figure 3, it can be observed that France continues to report the highest number of new covid–19 cases after the lockdown is lifted. During the second phase of pandemic, countries have largely avoided imposing nationwide lockdowns and are instead relying on partial lockdowns and targeted restrictions on movement in hot spots.

**Fig. 2:**
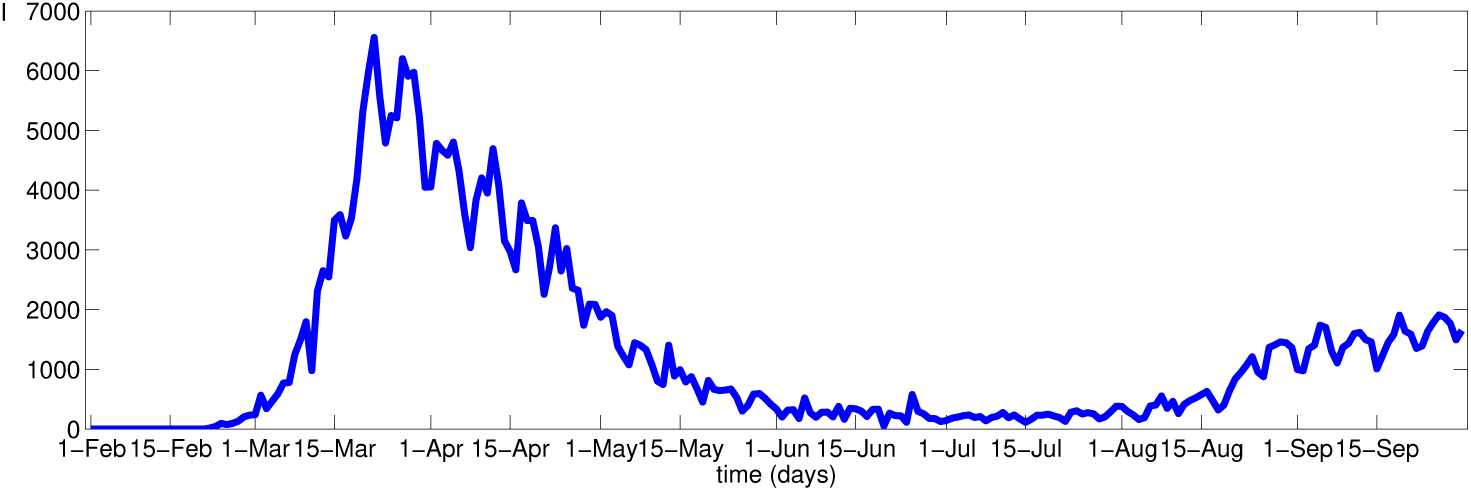
Daily reported COVID–19 cases from 31^st^ January 2020 to 29^th^ September 2020, Italy (Source: Worldometer)

**Fig. 3:**
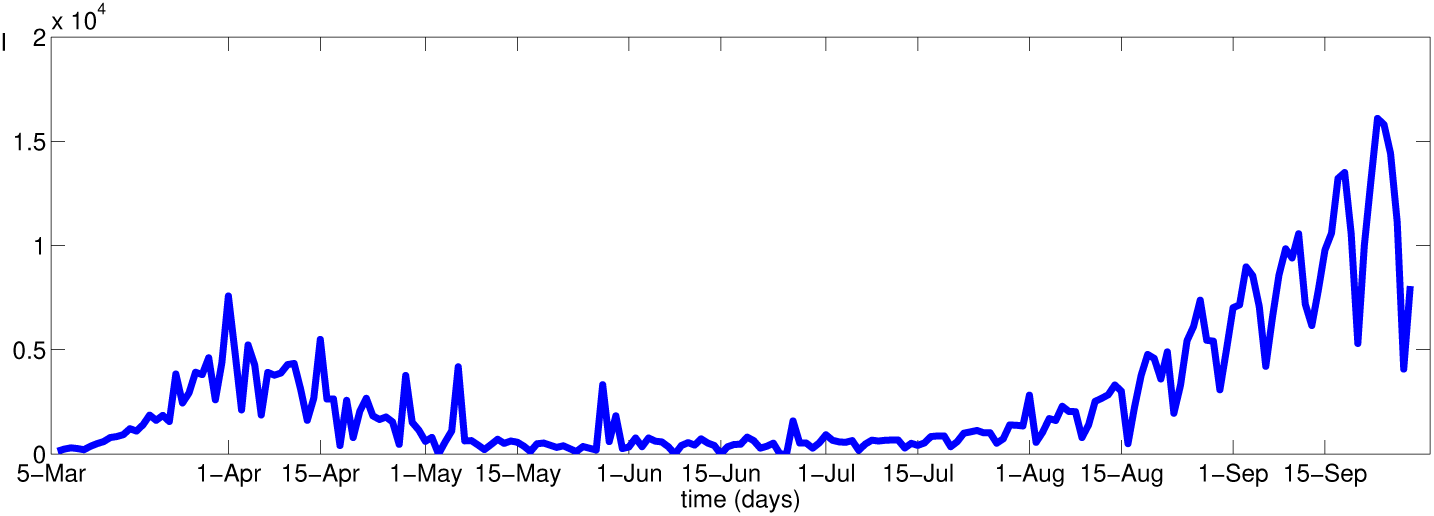
Daily reported COVID–19 cases from 5^th^ March 2020 to 29^th^ September 2020, France (Source: Worldometer)

In this context, it is important to seek what preventive measures are most appropriate to keep the pandemic controlled, while causing the minimum damage to the economy of the country. In other words, it is important to design successful partial lockdown strategies, or, lockdown relaxation strategies, by taking into account the transmission of the disease when travel is permitted and also the economic concerns. We propose a region–based lockdown relaxation strategy, which shares certain similarities with what was implemented during the lockdown period in several countries. Accordingly, we propose to determine the extent of lockdown a region must undergo, in order to limit the number of covid–19 patients to a number that can be provided necessary healthcare facilities using the current capacity in the country. It is a well known fact that the death rate from covid–19 rapidly increases when a country is unable to provide intensive care unit (ICU) beds and other necessary health facilities to the patients. Therefore, we assume that the country can reinforce a certain number of patients, during a given time. Also we consider the transmission dynamics of covid–19 from available data. Thus, we build an optimization model which determines the extent of lockdown that must be imposed on a region during the post–lockdown period.

In order to obtain computational results from the optimization model developed thus, we use the covid-19 data in Sri Lanka, where strict and mild lockdowns were implemented at several occasions. In order to enhance the computational experience, we describe the covid–19 situation in Sri Lanka in section 2. In section 3 we forecast the post–lockdown transmission of covid–19 using compartment models in epidemiology, by incorporating the inter–regional mobility factor. In section 4 we develop the optimization model. Section 5 includes our computational techniques and section 6 the interpretation and the discussion of the results we obtained. We conclude the paper in section 7.

## 2 Lockdown strategies in Sri Lanka

Sri Lanka reported the first covid–19 case in a Chinese tourist on 27^th^ January 2020 and subsequently in a local person on 11^th^ March, 2020. Aimed at controlling the pandemic, the government of Sri Lanka implemented several strategies, of which reducing human mobility was the most prominent one [8]. A strict strategy of lockdown was enforced together with other preventive measures including case detection, identification of contacts, quarantine, travel restrictions and isolation of small villages as well. Despite those preventive measures, the pandemic continues to threaten the public with daily reported cases. As seen from Figure 4, the reported covid–19 cases in Sri Lanka is still on the rise thus the country is still at risk. Further, Sri Lanka has experienced fluctuating doubling times below 70, as depicted in Figure 5, which questions the appropriateness of relaxations. On the other hand, the impact of the preventive measures to the economy of Sri Lanka was significant. Sri Lankan economy was slowly recovering from the Easter Sunday attacks in April 2019, and the Central Bank of Sri Lanka (CBSL) was expecting an economic growth of 4.5–5% together with the political stability after the recently held presidential and general elections. In this context, a continuing lockdown was regarded as quite impossible for the small island nation. Therefore, the government of Sri Lanka declared that the activities of the country will be restarted from 11^th^ May onwards, subject to several restrictions, aimed at a resuming to ordinary life gradually. The restrictions include social distancing, limitation of the workforce at workplaces etc. Also the nationwide curfew imposed for 52 days was lifted for many regions, except for several regions identified as high–risk zones. A limited number of citizens in high–risk zones were allowed to travel, based on the last digit of their national identity card numbers. Accordingly, specific days are prescribed for the individuals who possess national identity card numbers ending up at different digits, intending to reduce the human movement inside those zones by more than 80%. In addition, one third of the workforce in state institutions in several areas were required to report to work. Also the public transport services were made allowed with strict restrictions on the numbers of passengers. Thus, the country had undergone a transition from strict lockdowns towards partial lockdowns.

**Fig. 4:**
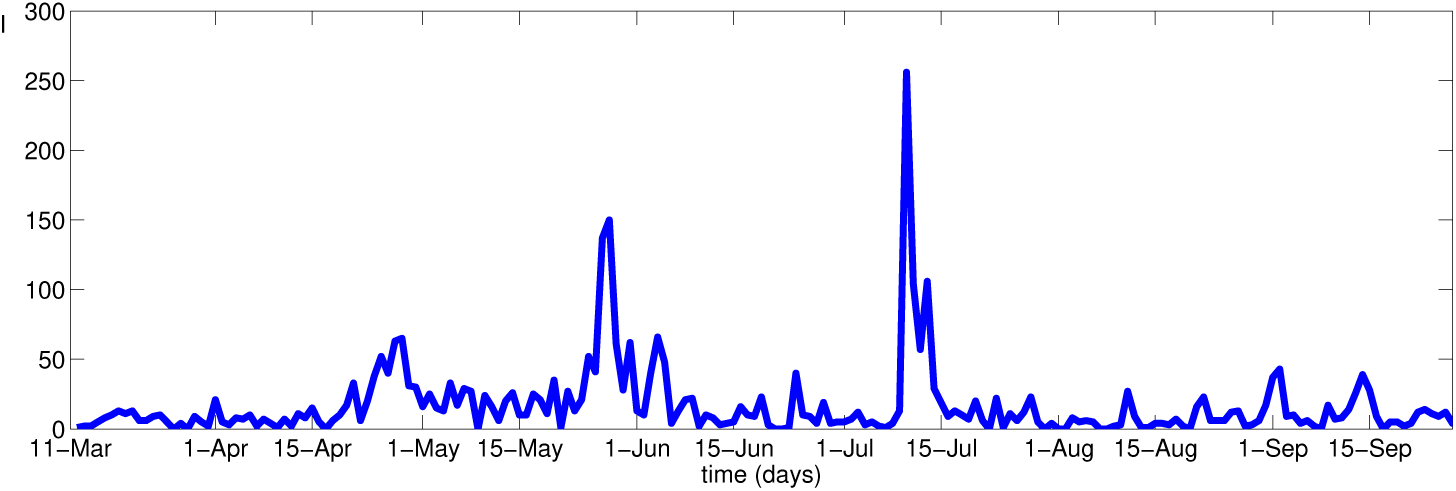
Daily reported COVID–19 cases from 11^th^ March 2020 to 29^th^ September 2020, Sri Lanka (Source: Epidemiology Unit, Ministry of Health, Sri Lanka)

**Fig. 5:**
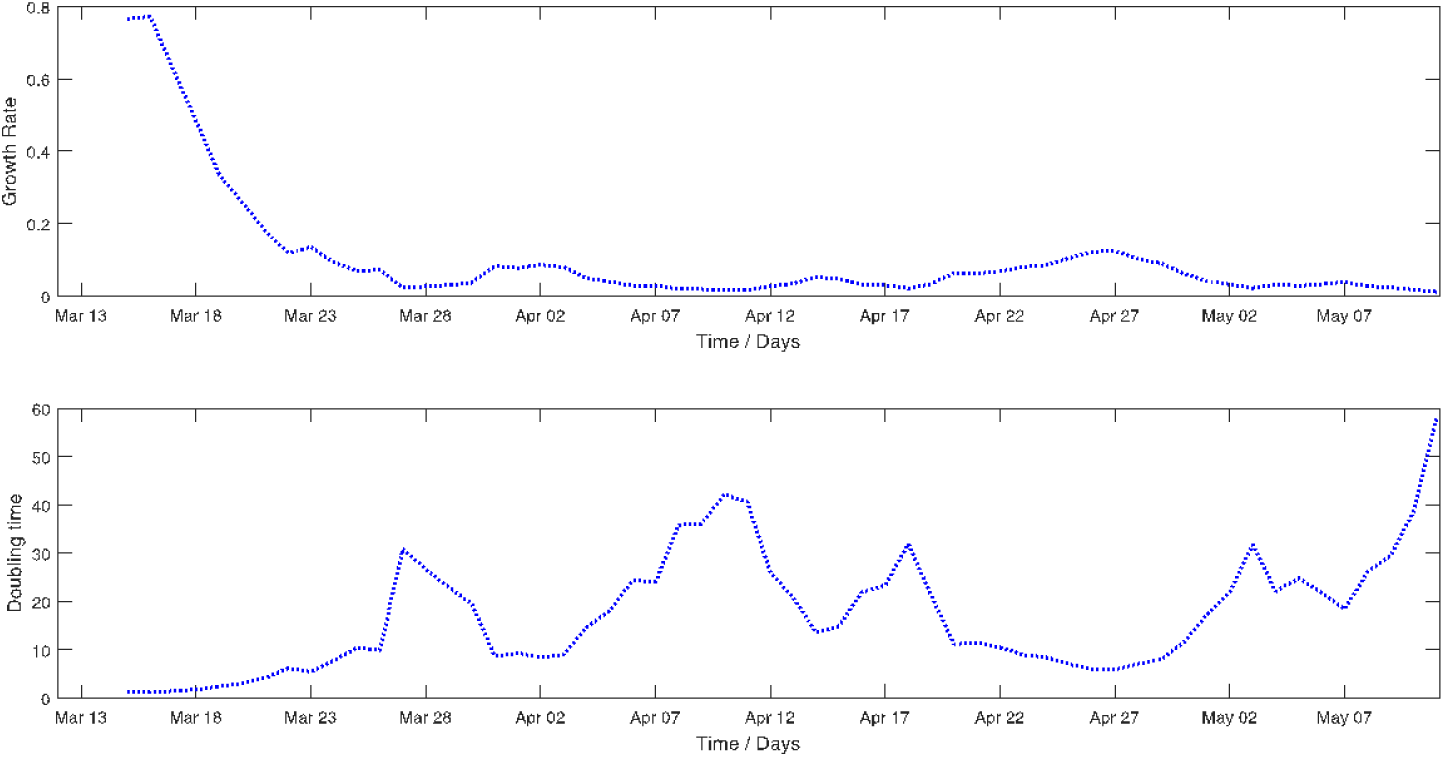
Growth rate and doubling time for covid-19 cases from 14^th^ March to 11^th^ May, Sri Lanka (Source: Epidemiology Unit, Ministry of Health, Sri Lanka)

It must be noted that even when the strict lockdowns were implemented in the country’s most populous Western province and the high–risk North Western province, partial lockdowns were implemented in several other provinces. For instance, human mobility in the North Central province which contributes most to agriculture was least restricted, a decision which helped the minimally interrupted distribution of rice and vegetables to the regions on which strict curfew was imposed. Also, the manufacturing industries were permitted to operate to a certain extent, prioritizing food and beverages. This was done by allowing a certain percentage of workers at food manufacturing industry to go to work in some regions, subject to strict measures on distancing. Moreover, temporary relaxations of curfew were exercised in several regions at different occasions. Therefore, the country has already undergone partial lockdowns, where different regions were operated to different extents, eventually contributing to the sustainment of the nation’s economy during the lockdowns. It must be noted that the country underwent partial lockdowns at several times, most recently after the identification of the cluster that exceeded 1000 positive cases from an apparel factory in the Western Province in early October 2020.

Nevertheless, the days of lockdown should not be seen as identical to the new post–lockdown period. A major distinction of the new period from the lockdown days is the inter–regional travel which was allowed from 11^th^ May onwards. Recall the major control measure for covid–19 was strictly restricting the human movement inside the country, its relaxation must have unforeseen consequences. Several previous works on epidemiology have pointed out the significance of human mobility to the transmission of covid–19 [9,24]. Therefore, the relaxation of travel restrictions must put a different complexion on the matter, making the post–lockdown period significantly different to the lockdown period.

## 3 Post–lockdown disease transmission

In order to model the disease transmission in the country that undergoes a lockdown (or a relaxation), we adopt the SIR (susceptible–infectious–recovered) model for epidemic transmission, which is the most frequently used compartment model to forecast an epidemic. This model captures the transmission dynamics of diseases of which the infection confers permanent immunity. The population (*N*) is divided into the three disjoint classes, namely, susceptible (*S*), infectious (*I*) and recovered (*R*). Once the susceptible individuals become infected with the disease, they move to the infectious class. The infected persons move to the recovered class when they get recovered from the disease and a person in the recovered class is assumed to have permanent immunity. Let *β* denote the transmission rate from infected individuals to susceptible individuals and *γ* denote the recovery rate of infected individuals. Then, the timely variation of the compartments is described by the following set of differential equations:

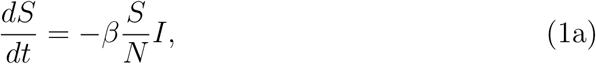

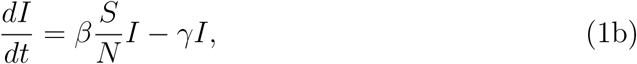

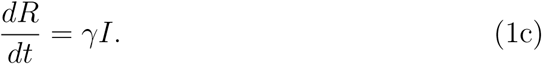

Several researchers have used the SIR model to forecast the covid–19 incidence [3,5,23]. The SIR model in its original form is however not very helpful for our purpose. Recall inter–provincial travel is the main characteristic which distinguishes the post–lockdown period in our context, in order to forecast the transmission during that period, it is essential to incorporate the travel component to the conventional SIR model. Hence, we modify the SIR as follows.

Let *w*_*ij*_ denote the percentage of daily travels from *i*th province to the *j*th province and degree of social distancing respectively. Let *x*_*i*_ denote the lockdown relaxation percentage of the *i* th province. Then the susceptible (*S*_*i*_), infectious (*I*_*i*_), recovered (*R*_*i*_) populations in *i*th province can be described by the following set of modified differential equations.

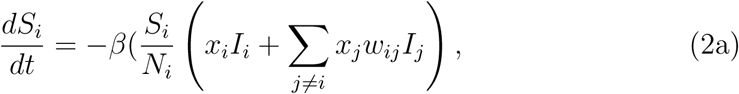

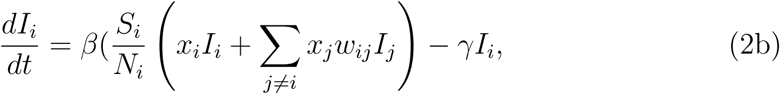

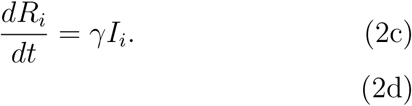

Following the works by [17,18], we define the expected number of transmissions an individual has received by time *t* as:

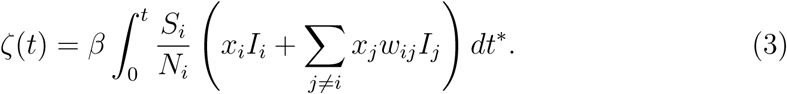

Notice that

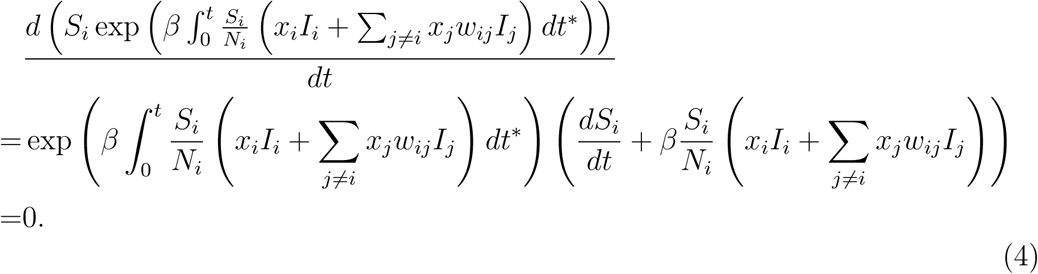

Thus, it follows that

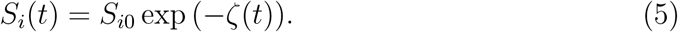

Further, by integration Equation 3, it is possible to show that

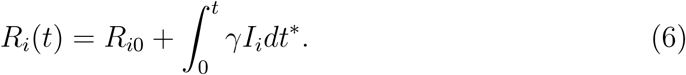

By substituting *S*_*i*_(*t*) and *R*_*i*_(*t*) into *N*_*i*_(*t*) = *S*_*i*_(*t*) + *I*_*i*_(*t*) + *R*_*i*_(*t*), infected human population at time *t* according to our model can be expressed as:

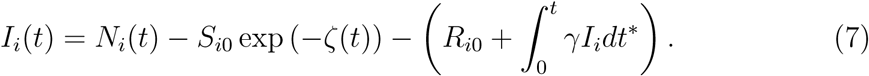

## 4 Proposed optimization model

Several works have investigated the critical subject of intensive care management during covid–19 pandemic with different opinions [7,22,28], which we do not wish to explore in this work. Determining who would be provided with ICU facilities is another decision problem we do not wish to address in this work. Instead, we consider a situation where all patients are provided ICU beds. If the epidemiological recommendations suggest otherwise, this assumption can be readily relaxed and the relevant term can be replaced by the percentage of the infected population who are facilitated with ICU beds. Accordingly, we state the condition that the number of covid–19 patients in the post–lockdown era, which is given by Equation 7 must not exceed the number of ICU beds.

Once the transmission of the disease to different provinces during the post– lockdown period is formulated, it is now important to examine the contribution of these provinces to the economy. Recall the decisions on curfew were made at different notes on different regions during the 52 days of lockdown, a primary intention was sustaining the nationwide economy by considering the economic contribution from regions. Our model is also based on the regular (or the pre–lockdown) economic contribution by different provinces. For this purpose, we use the data in Sri Lanka, which is given in Table 1.

**Table 1:** Agriculture and industrial contribution from each province to the economy of Sri Lanka (Source: Economic Statistics of Sri Lanka, Department of Census and Statistics, Sri Lanka)

| Province | Agricultural contribution (%) | Industrial contribution (%) |
| --- | --- | --- |
| Western | 2.03281 | 13.79459 |
| Central | 12.05421 | 11.08724 |
| Southern | 10.41369 | 11.73184 |
| Northern | 10.23538 | 10.39966 |
| Eastern | 9.700428 | 10.78642 |
| North Western | 9.379458 | 14.00945 |
| North Central | 15.29957 | 8.293941 |
| Uva | 18.61626 | 6.574989 |
| Sabaragamuwa | 12.26819 | 13.32187 |

Since our intention is determining to what extent a province would operate during the post–lockdown era, the decision variable in our optimization model must be the extent of lockdown relaxation of the *i*th province, given as a percentage, symbolized by *x*_*i*_ in section 3. Then the inputs follow as given below.

*m* : Number of provinces

*p*_*i*_ : Economic productivity of the *i*th province

*w*_*ji*_ : Human mobility between the *i*th and the *j*th provinces

*β* : Transmission rate of covid–19

*γ* : Recovery rate of covid–19

*N*_*i*_ : Population of the *i*th province

*M* : Number of ICU beds available in the country

*S*_*i*0_ : Initial susceptible population in the *i*th province

*R*_*i*0_ : Initial recovered population in the *i*th province

Now, the relevant optimization problem can be expressed as follows.

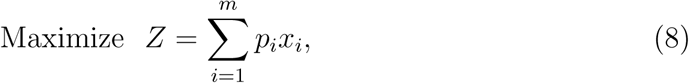

subject to

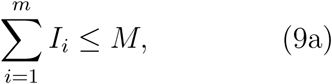

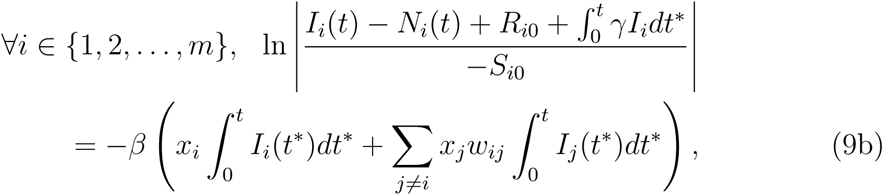

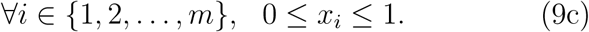

The objective function 8 maximizes the total contribution to the economy by all provinces. Constraint 9a assures that the number of total infected persons within the relevant period of time does not exceed the number of ICU beds in the country. The inter–regional transmission of the disease as obtained by applying the SIR model is given by the set of constraints in 9b. Finally, constraints in 9c assure that the percentage of lockdown of any province must be between 0% and 100%.

## 5 Solution technique

In order to solve the non–linear optimization problem given by Equations 8 and 9, we consider a particular characteristic of the objective function 8 and constraints 9a, 9c. That is, despite being multivariate functions, all these are expressible as sums of single–variabled functions. Also the constraint 9c is readily transformable to this form by substituting 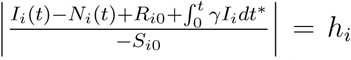 and *f*_*i*_ = exp −*h*_*i*_. Therefore, we can restate our optimization problem as follows.

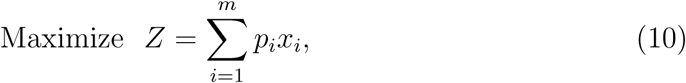

subject to

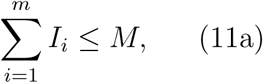

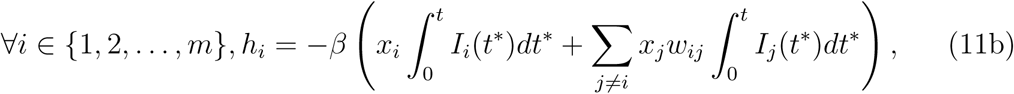

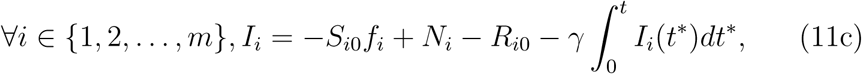

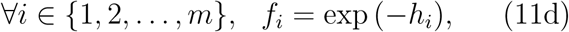

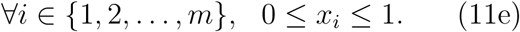

This reformulation given by Equations 10 and 11 motivates us to adopt the technique of *separable programming*. This technique was first introduced in [6] for constrained optimization of non–linear convex functions, whenever these functions are separable; that is, expressible as sums of functions of single variable. Since its inception, *separable programming* has been a very useful optimization technique, with applications to several real–world problems including agricultural planning [27], linear complementarity problem [4], Newsboy problem [1] and demand allocation [11]. The main tool in separable programming is, replacing the non–linear functions in the optimization problem by piecewise linear approximations.

Notice that Equation 11d in our formulation is non–linear and hence piecewise linearization is required for the function *f*_*i*_. We divide the domain of *f*_*i*_, that is [0, *D*_*i*_], into *n* subdivisions, each of length *d* by defining *h*_*ik*_ as follows, where, *D*_*i*_ is the maximum value of *h*_*i*_.

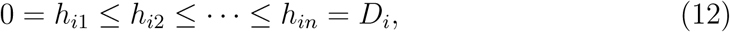

where

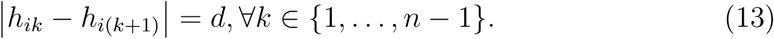

Then any point *h*_*i*_ in the interval [0, *D*_*i*_] can be uniquely expressed as:

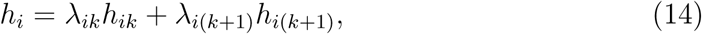

where

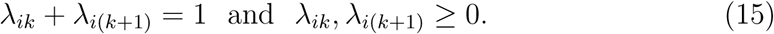

Now the piecewise linear approximation to *f*_*i*_ is expressible as:

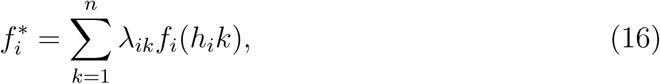

where

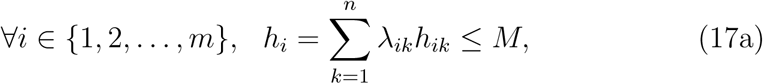

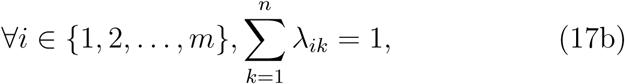

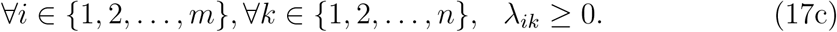

with the additional restriction at most two adjacent *λ*_*ik*_’s are positive.

Replacing the non–linear functions *f*_*i*_ in Equation 11d by linear approximations in Equation 17, our problem can be restated again as follows:

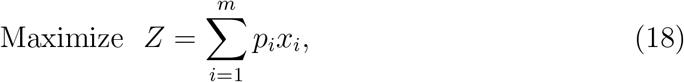

subject to

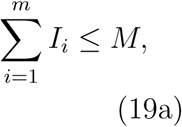

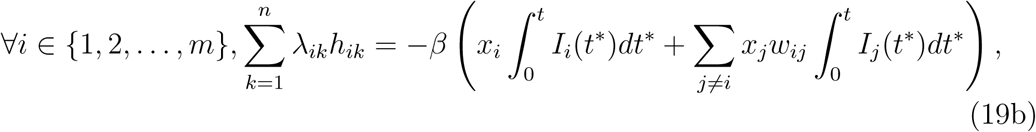

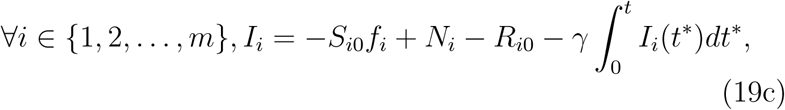

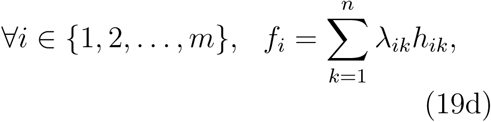

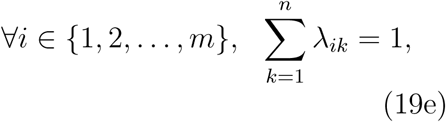

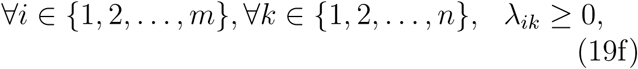

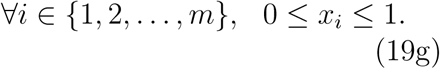

with the additional restriction that at most two adjacent *λ*_*ik*_’s are positive.

Except for the additional restriction on adjacency, the approximated problem given by Equations 18 and 19 is a linear program, readily solvable by the simplex method. It is a standard fact in separable programming that, in case of maximization, if the approximated objective function is concave and each piecewise linear constraint is convex, then the solution of the linearly approximated formulation without the additional restriction is feasible to the original problem [12,25]. From this, the computational hardness implied by the non–linearity could be readily overcome, and the problem becomes efficiently solvable.

## 6 Results

Based on the information available on covid–19 pandemic in Sri Lanka, we made substitutions of numerical values to the inputs. First, Sri Lanka currently has approximately 500 ICU beds, and the authorities have recently declared their willingness to increase this to 1000. Therefore, the input *M* was set to 1000 in our primary computational experiment. Further, the covid–19 incidence by province as per 11^th^ May were taken as the initial conditions. This selection of initial condition is a realistic choice required by the model, as it is the day the country started undergoing relaxations. Provincial economic contribution was substituted as in Table 1. The duration the model applied was taken as one week from the day the relaxations started.

The global optimum to the problem achieved through separable programming, applied to these data, provided the best relaxation percentages of the provinces, as given by Table 2 and visualized in Figure 6. There are several interesting and counterintuitive implications. According to these results, no relaxation must be done on the lockdown of the Western province; moreover, it is not the only province which must be kept at strict lockdowns; also the Central and the North Western provinces must undergo strict lockdowns. On the other hand, total relaxation is possible for five provinces, namely, Southern, Northern, Eastern, North Central and Uva. Recall the North Central province, the country’s agricultural hub, underwent least lockdown measures till 11^th^ May to assure the people are provided with rice and vegetables, our results too indicate that the agricultural farming can be restarted in this province and its economic centres could be kept open during the post–lockdown period.

**Table 2:** Optimal lockdown relaxations of provinces as percentages (Number of ICU beds =1000)

| Province | $x_i$ |
| --- | --- |
| Western | 0 |
| Central | 0 |
| Southern | 100 |
| Northern | 100 |
| Eastern | 100 |
| North Western | 0 |
| North Central | 100 |
| Uva | 100 |
| Sabaragamuwa | 57 |

**Fig. 6:**
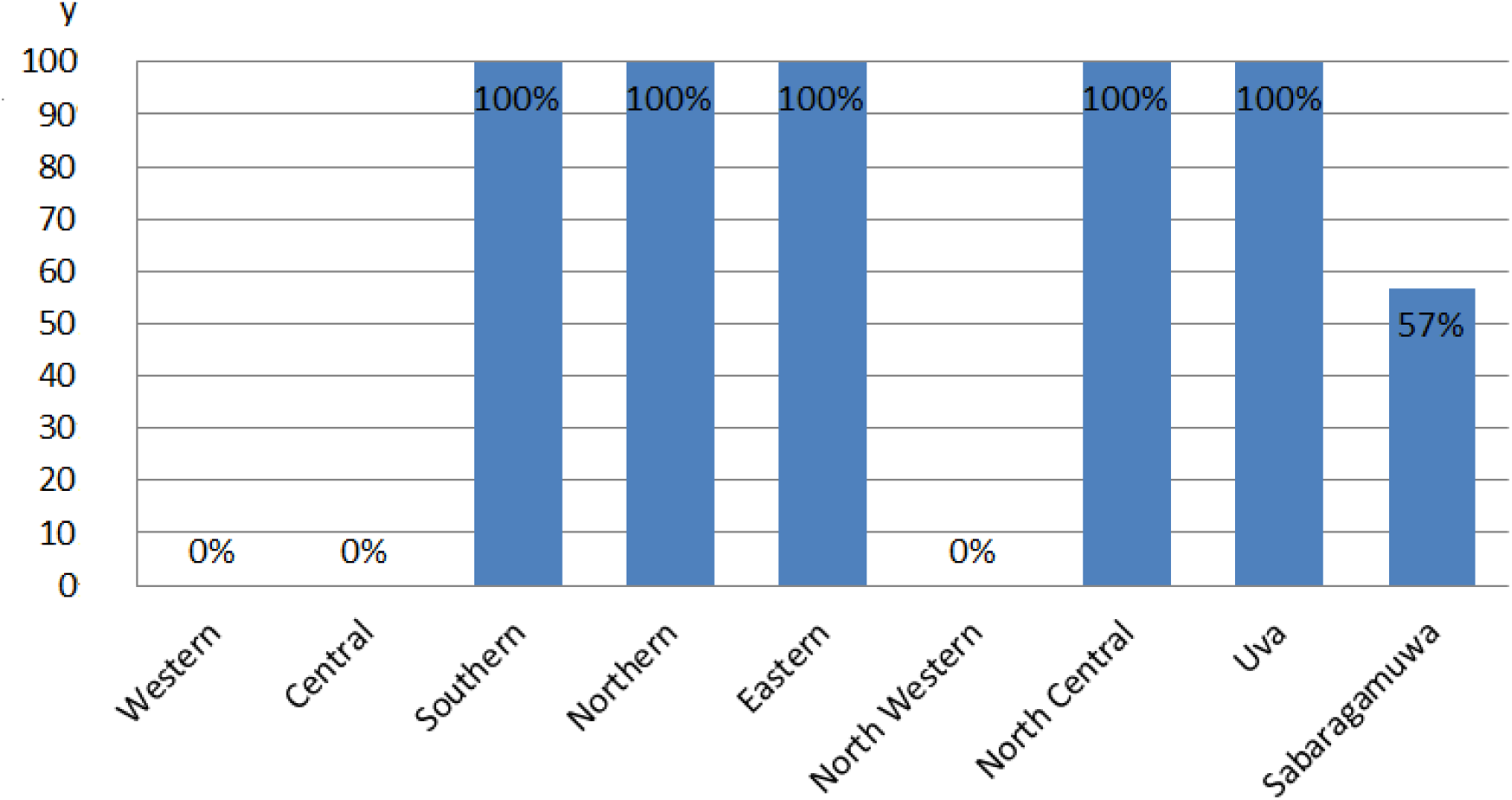
Optimal results for lockdown relaxation (y - Relaxation level as a percentage, Number of ICU beds =1000)

Our secondary computational experiment was on investigating the scenario when different numbers of ICU beds are available. The results are shown in Figure 7. Recall Sri Lanka currently has 500 ICU beds in state hospitals, the results indicate that relaxations must be applied only to three provinces, namely, Southern, Eastern and North Central. The Northern province also could undergo a minor relaxation, if the number is increased to 750, in addition to the significant fact that Uva province might undergo a major relaxation. The lockdown imposed on the Central province can be partly relaxed at a stage when the number of ICU beds is in between 1000 and 1250. Sabaragamuwa province could undergo a partial relaxation when this number is between 750 and 100. Western and North Western provinces must remain unrelaxed even if the number is increased to 2000.

**Fig. 7:**
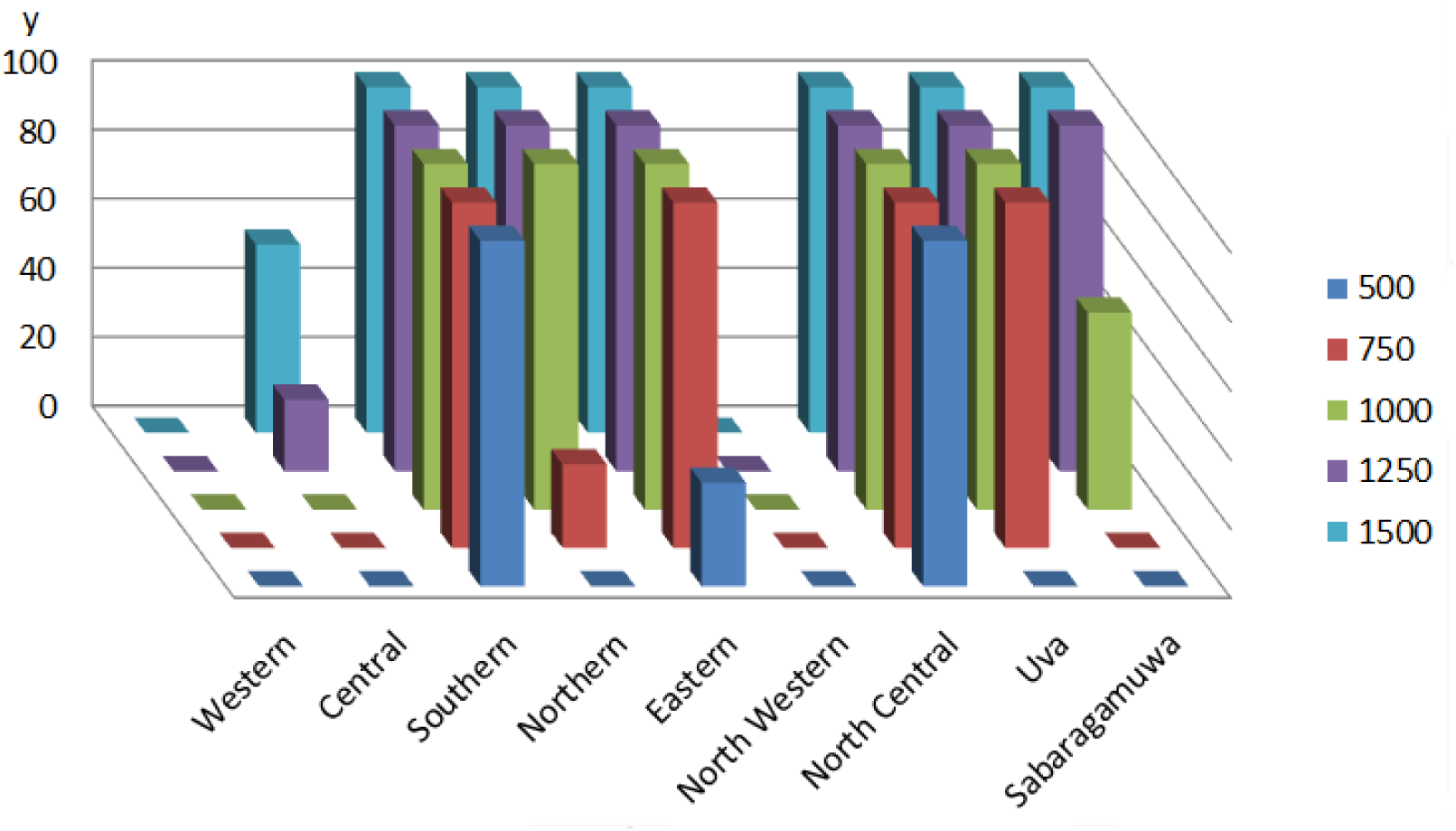
Variation of the optimal lockdown relaxation levels of provinces (as percentages) with the total number of ICU beds available in the country

## 7 Conclusion

Human mobility is a critical factor in transmitting communicable diseases. Accordingly, lockdowns with strict travel restrictions were implemented in several countries to fight against the covid–19 pandemic. A nationwide curfew was implemented in several countries including Sri Lanka and India, which had a significant impact on the economy. Since minimizing both covid–19 incidence and economic effects are two conflicting goals, resuming to economic activities is inescapable. Therefore, countries consider partial lockdowns, aimed at a gradual transition to ordinary life. Instead of ad–hoc lockdown or relaxation strategies, in this work we proposed a systematic lockdown relaxation strategy that can help in achieving the two conflicting goals.

Considering the potential disease transmission during the post–lockdown period, as given by the epidemic models, we proposed an optimization model, from which the optimal region–based lockdown strategies were determined, while confining the covid–19 incidence to a number that is endurable to the country and minimizing the damage to its economy. In particular, we proposed to determine the extent of lockdown relaxation for each region such that all covid–19 patients could be provided ICU facilities and the contribution to economy by all provinces is maximized. Since the resulting optimization problem turned out to be non–linear and several of its functions were expressible as sums of single–variabled functions, we adopted the method of separable programming to generate solutions for the relevant data sets in Sri Lanka. Accordingly, we converted the non–linear functions to piecewise linear approximations and found the global optimum.

In a more realistic setting, other constraints must be added to our model. For instance, it was mentioned in section 2 that during the lockdown period, the North Central province which acts as the agricultural hub in Sri Lanka underwent least lockdown restrictions. If all agrarian activities must be continued in the post– lockdown period, the relevant constraint can be readily incorporated into our model, as a lower bound to the relaxation of this province. That adds another linear, single– variabled and convex constraint which does not change the solution criterion. However, if the government is looking forward to running different specific industries in different regions, our model needs further modifications. In that case, a decision variable could be indexed by two indices, one for the province and the other for the industry. Further, our model can be applied at any moment in the post–lockdown period by substituting relevant initial conditions. In addition, even if travel restrictions were reimposed on certain roads, the relevant input can be changed and the updated optimal lockdown levels could be determined by minimum modifications. Due to concavity and convexity of the objective functions and constrains, it is efficiently solvable, even if the inter–provincial mobility information were replaced by a subtler dataset such as inter–district mobility; which could be a significant improvement of our model in light of accuracy. Accordingly, our optimization model can be improved further to help the decision making process in sustaining the economy in post–lockdown times, preventing the excessive transmission of covid–19. It would be an interesting future research task to formulate an analogous optimization problem, in which the factors that integrate the individual economic contribution by provinces are included.

## 8 Data Availability

COVID–19 data can be retrieved via Epidemiology Unit, Ministry of Health, Sri Lanka, Available at:

*http://www.epid.gov.lk/web/index.php?option=comcontent&view=article&id=225&Itemid=518&lang=en* and mobility data can be retrieved via National Transport Commission, Sri Lanka, Available at: *https://www.ntc.gov.lk/corporate/pdf/statistics%202015.pdf*

